# Modeling the dynamics of COVID-19 using Q-SEIR model with age-stratified infection probability

**DOI:** 10.1101/2020.05.20.20095406

**Authors:** Joshua Frankie Rayo, Romulo de Castro, Jesus Emmanuel Sevilleja, Vena Pearl Boñgolan

## Abstract

We explore the advantage of age-stratifying the population as an improvement on the quarantine-modified SEIR model. We hypothesize that this would project lower cases of infection for the Philippines because of our country’s low median age. We introduce the variable *U* that is multiplied to the incubation rate *σ* when exposed individuals become infected. *U* is the dot product of the proxy infection probabilities stratified per age group (*F*) and the population stratified per age group (*P*) divided by the total population, similar to calculating mathematical expectation. Proxies were taken from two data sets: Hubei, China with a calculated value of *U_CHN_* = 0.4447 and Quezon City, Philippines with *U_QC_* = 0.5074. When the majority age group, represented by the median age, is far from the age group with the highest number of infections the number of infected individuals decreases and produces a delayed peaking effect. This new method gives a much lower estimate on peak number of infected cases by 65.2% compared with age-stratification alone; and by 75.2% compared with Q-SEIR alone.

## Introduction

As COVID-19 rages around the world, modeling of the epidemic must be continuously validated against the actual number of cases to help decision makers plan healthcare resource allocations. This scenario underscores the importance of massive, inclusive diagnostic testing to get to the real number of cases in a locality.

The Q-SEIR model [1] is a compartmental model that estimates the number of susceptible (*S*), exposed (*E*), infectious (*I*) and removed (*R*) individuals during an epidemic in a community (*N*) with quarantine factor (*Q*). The dynamics of this epidemic can be described by a system of differential equations

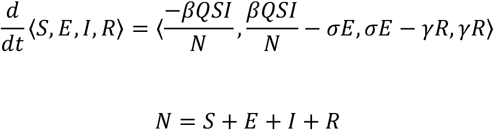

Parameters needed to run the model include: *t* as unit of time, usually in days and starts at *t* = 0, *β* is the transmission rate (number of individuals who contract the disease per unit time), *σ* is incubation rate for the symptoms of the disease to manifest on an individual (expressed as 1 person per units of time) and *γ* is removal rate (expressed as 1 person per units of time). This model does not consider vaccination and ignores the population’s vital statistics since the epidemic may not linger long enough for natural births and deaths to be influential. It also assumes removed as either recovery with permanent immunity or death.

*Q*(*t*) is a parameter that represents the effectiveness of quarantine, which can change over time. A value of 1 means that there is no quarantine and we regain the original SEIR model, while a value of 0.4 means that it is 60% effective. Implementing quarantine effectively reduces disease transmission: less people will be exposed consequently lowering the number of infected individuals, delaying the peak, and flattening the epidemic curve as expected. Using the Age Stratification method, for Quezon City, the projected peak number of infected cases is 9.95% of the total population, while Q-SEIR projected 13.99% with *Q*(*t*) = 0.4 [1].

The Q-SEIR model assumes a homogeneous population where all individuals have the same probability of becoming infected. However, in Hubei, China, the report [5] indicates that number of infected individuals disproportionately occurs in higher age groups. We treat the age-stratified occurrences in Hubei as good proxies for the infection probabilities per age group in Quezon City, Philippines. Thus, we now combine the Q-SEIR model and the presumed age-stratified infection probability. We hypothesize that this would project a lower number of infections for Quezon City because of the Philippines’ younger population. The number of infected cases at the peak and the time of peaking using this model (Age-Stratified Q-SEIR, ASQ) will be compared with Q-SEIR.

## Materials and Methods

We assumed that *β*, *σ* and *γ* parameters are immutable properties of the virus-host interaction therefore they remain constant throughout the simulation. Two variables (*Q* and *U)* were introduced to incorporate the quarantine policy and age-stratification of infection probability, respectively. The system is globally stable, so it was solved using first-order forward Euler method. *N* is conserved on each iteration, 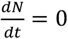. A Microsoft Excel worksheet was used in entering the necessary parameters, implementing the numerical solution, and displaying the simulation results. The following parameter values were assigned for the base configuration:

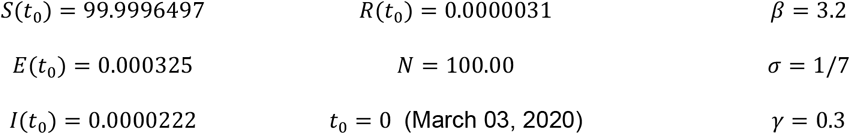

Simulation results on different scenarios are compared from this base configuration.

## Results and Discussion

Fig 1 and Table 1 show simulation results on Q-SEIR for Quezon City where *Q*(*t*) = 1 from day 0 to 13 and *Q*(*t*) = 0.2 (80% successful quarantine) from day 14 onwards – the initial confirmed cases appeared on March 03 and the quarantine was imposed on March 16. This long-term scenario seems to have recreated *herd immunity*, protecting 16.44% of the susceptible population after 83.56% of the population was removed. Quarantine policy *Q*(*t*) = 0.2 decreased the peak number of infected individuals, from 13.99% [1] to 5.79%.

**Fig 1.**
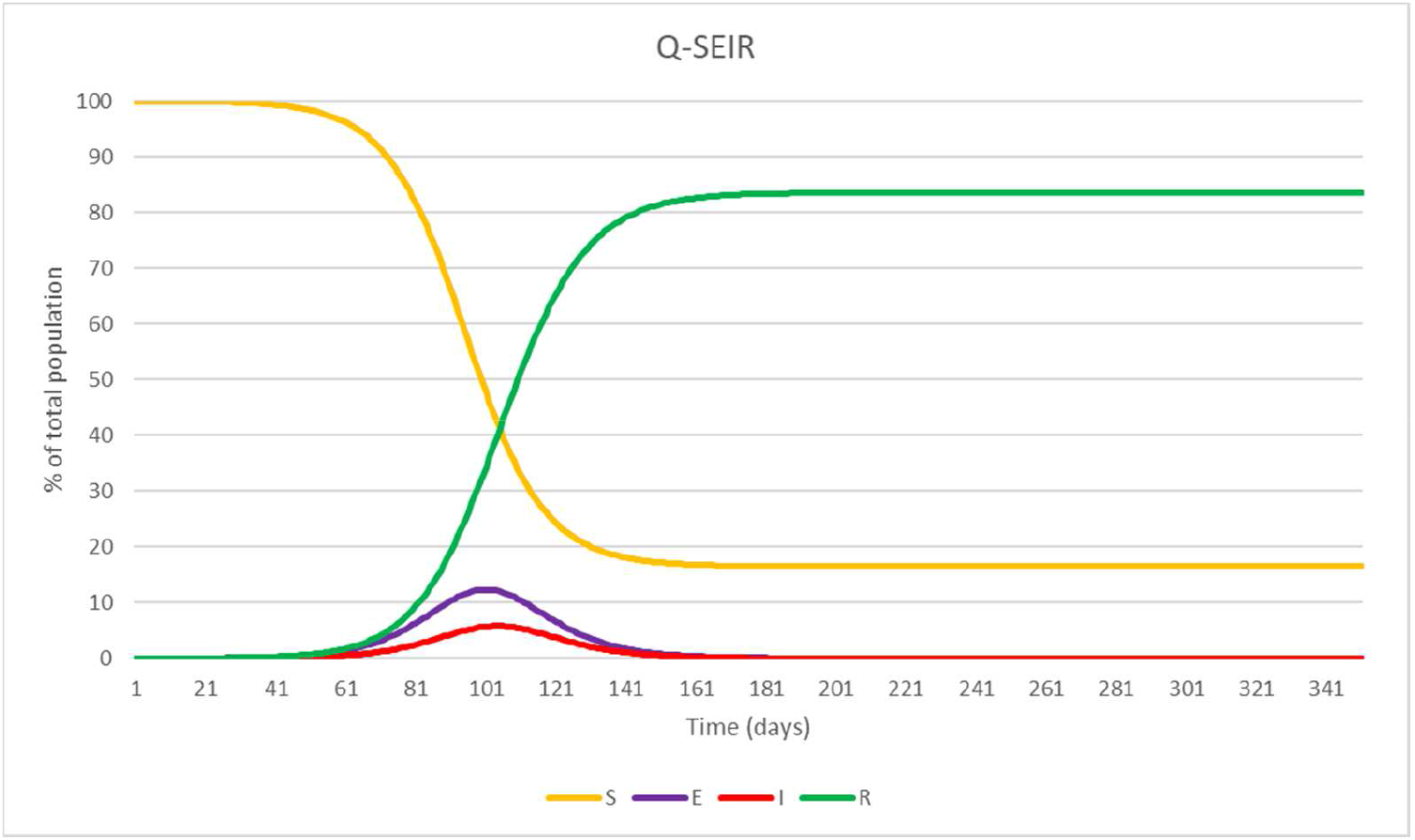
Q-SEIR model simulation displaying a herd immunity effect after 161 days of continuous quarantine. *Q*(*t*) = 1 from day 0 to 13 and *Q*(*t*) = 0.2 from day 14 onwards.

**Table 1.** Peak times and values from Q-SEIR model with stricter quarantine.

| Findings | Time (days) | Date | Percentage |
| --- | --- | --- | --- |
| Peak exposed | 100 | 28/06/2020 | 12.3241 |
| Peak infected | 103 | 01/07/2020 | 5.7914 |

An early report [5] showed that COVID-19 has a higher infection occurrence among the older age groups and those with comorbidities. As an improvement to the Q-SEIR, an age-stratification component was added (we now call as ASQ) where the relative proportion of infected cases per age group proxies the infection probability. We only modified the term involving the transition from *E* to *I*. The probability of infection was represented by parameter *U*, such that 0 < *U*(*t*) ≤ 1, which is multiplied to *σ*. The effect is that the incubation time, averaged across the population over all age stratifications, increases. When *Uσ < γ*, patients are being discharged from hospitals faster than being admitted.

Without loss of generality, we use nine bins to group individuals by 10-year age intervals (0-9 years old, 10-19 years old, …, 70-79 years old to 80 years old and above). Let *P* = {*P*_1_, …, *P*_9_} be the number of individuals per age group. Similarly, we define *F* = {*F*_1_, …, *F*_9_} as the number of infected individuals per age group (proxy for infection probability).

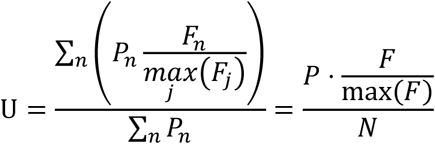

The quantity *F*/max (*F*) represents relative infection probability per age group, with computed values on Table 2. This is the age groups’ presumed infection probability relative to the most vulnerable age group. We define the most vulnerable age group as the age group with the greatest number of infected individuals. In our simulation, exposed individuals from the most vulnerable age group (*F*/max (*F*) = 1), become infected after the specified incubation time while individuals in other age groups (*F*/max (*F*) < 1) effectively extend their incubation time. Table 2 shows values for *P, F* and *F*/max (*F*) for Quezon City [2] and Hubei [5]. Quezon City reported 1,294 cases as of May 5, 2020 and Hubei reported 44,672 cases as of February 11, 2020. Fig 2 shows that they tend to have a normal distribution, a consequence of the Central Limit Theorem. It is observed that the most vulnerable age group in Quezon City (30-39 years old) is younger than Hubei (50-59 years old). We posit that this is a direct effect related to the Philippines’ young age [1, see also Kenya vs. Japan comparison].

**Table 2.** Relevant values for calculating the value of *U*. The second column contains the population of Quezon City by age group. The third and fourth columns contain the reported number of infected cases from Quezon City and Hubei by age group. The fifth and sixth columns are the normalized age-stratified infection probability proxies used to determine *U*.

| Age group<br>(years) | $P_{QC}$ | $F_{QC}$ | $F_{Hubei}$ | $F/\max(F)_{QC}$ | $F/\max(F)_{Hubei}$ |
| --- | --- | --- | --- | --- | --- |
| 0-9 | 573,845 | 12 | 416 | 0.0439 | 0.0415 |
| 10-19 | 612,165 | 20 | 549 | 0.0732 | 0.0548 |
| 20-29 | 659,804 | 166 | 3,619 | 0.6080 | 0.3616 |
| 30-39 | 512,688 | 273 | 7,600 | 1.0000 | 0.7594 |
| 40-49 | 390,696 | 192 | 8,571 | 0.7033 | 0.8564 |
| 50-59 | 275,683 | 231 | 10,008 | 0.8461 | 1.0000 |
| 60-69 | 142,813 | 243 | 8,583 | 0.8901 | 0.8576 |
| 70-79 | 52,146 | 120 | 3,918 | 0.4396 | 0.3914 |
| ≥80 | 21,870 | 37 | 1,408 | 0.1355 | 0.1407 |

**Fig 2.**
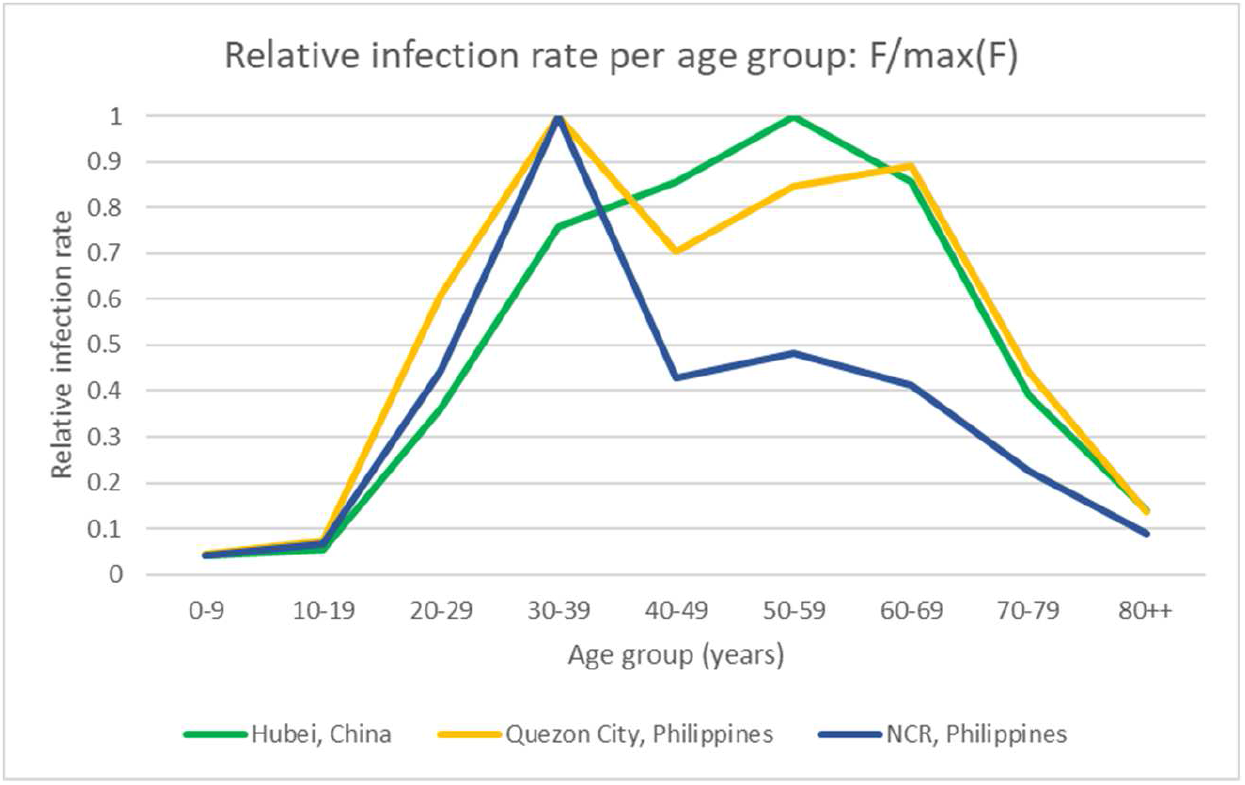
Distribution of relative infection probabilities from Hubei (as of February 11, 2020), Quezon City (as of May 05, 2020) and the National Capital Region (NCR), Philippines (as of May 14, 2020). Quezon City is a district of NCR. The most vulnerable age group (30-39 years old for both QC and NCR) is assigned a score of 1.

The Philippines has a very young population with a median age of 25.2 years, lower than the world median age of 30.9 years and China’s 38.4 years [3]. The vulnerable age range of the Quezon City is 30-39 years old, while Hubei is 50-59 years old. The value of *U* is close to 1 if *P* and *F* has similar spread. Normalization relative to the age group with most infected cases show: *U_Hubei_* = 0.4447 and *U_QC_* = 0.5074. *U_QC_* may still change as the confirmed cases of the epidemic continue to be tallied.

Fig 3 and Table 3 shows a simulation on Age-stratified Q-SEIR (ASQ) model for Quezon City where *Q*(*t*) = 1 from day 0 to 13 and *Q*(*t*) = 0.2 from day 14 onwards with age-stratified infection probability from Quezon City as well.

**Fig 3.**
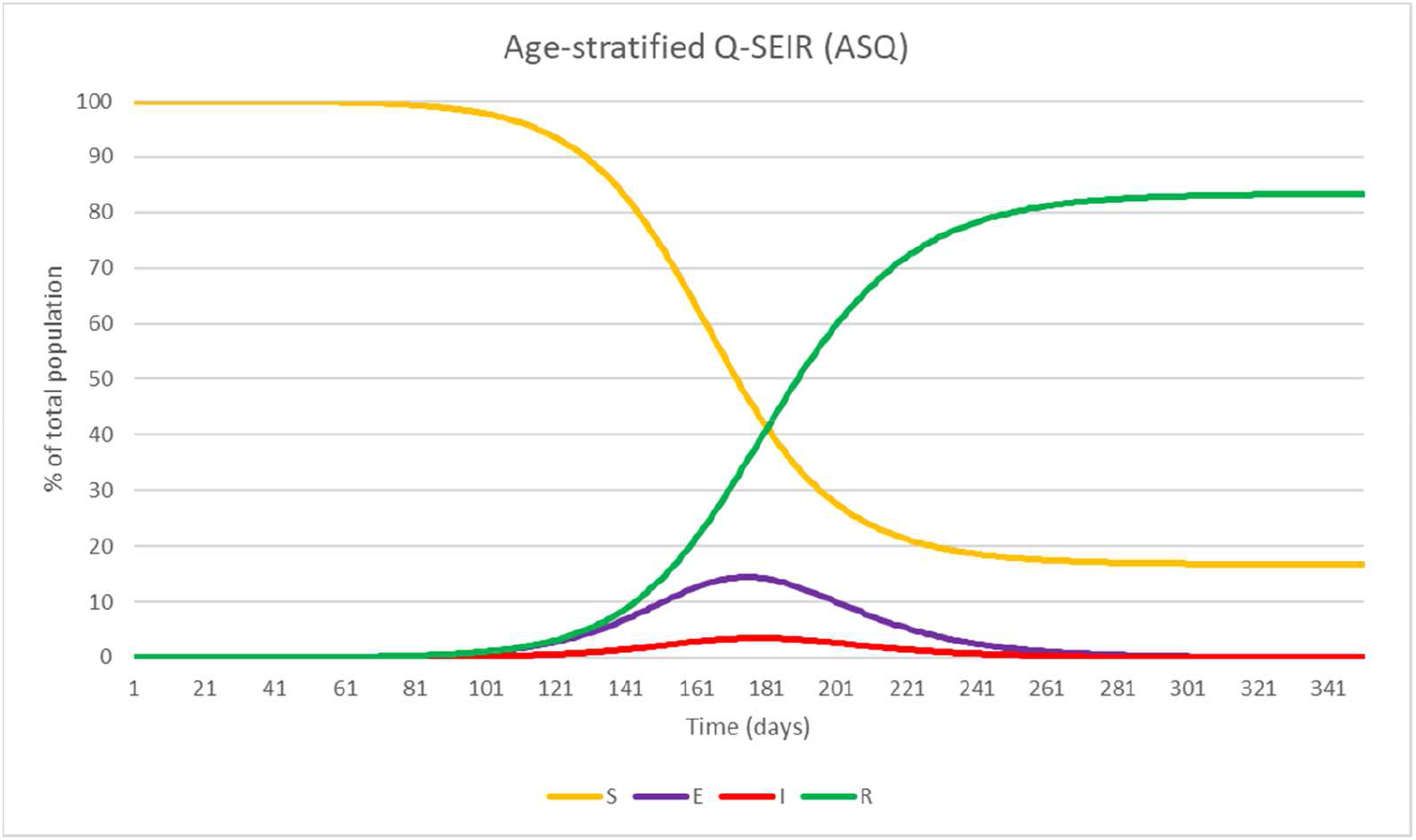
Age-stratified Q-SEIR (ASQ) model simulation. *Q*(*t*) = 1 from day 0 to 13 and *Q*(*t*) = 0.2 from day 14 onwards and *U*(*t*) = 0.5130.

**Table 3.** Peak times and values from Age-stratified Q-SEIR (ASQ) model simulation.

| Findings | Time (days) | Date | Percentage |
| --- | --- | --- | --- |
| Peak exposed | 174 | 10/09/2020 | 14.4199 |
| Peak infected | 178 | 14/09/2020 | 3.4673 |

Compared to Q-SEIR, the peak number of infected cases decreased from 5.79% to 3.47%. However, the peak number of exposed cases increased from 12.31% to 14.42%. Younger age seems to be protective (lowers infection) yet the higher proportion of younger individuals in a population increases exposure. This is the age-stratification effect on a young population if the older age groups are more vulnerable. This principle is well-illustrated by COVID-19 tests (Fig 4) and confirmed cases (Fig 5) from Hospital X, a government hospital from the Philippines, undisclosed for privacy concerns. A young population effectively prolongs the incubation period, very likely because of their better immune status and fewer risk factors such as diabetes and cardiovascular diseases.

**Fig 4.**
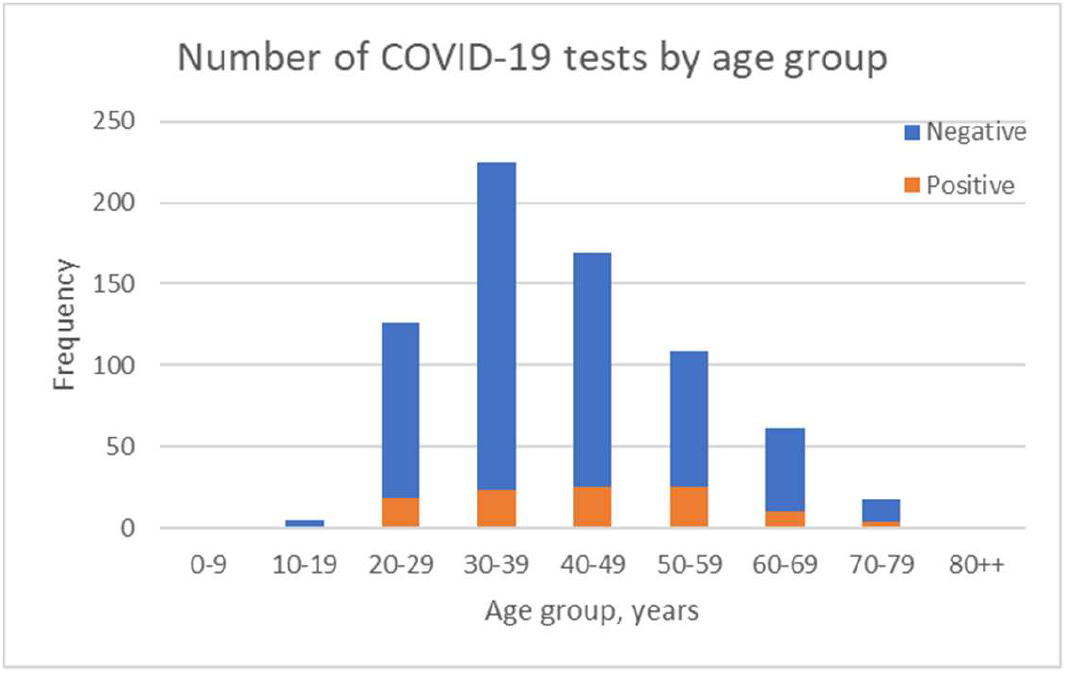
Results of COVID-19 testing implemented at Hospital X on May 05, 2020. There is a higher number of young individuals in this population which increases exposure.

**Fig 5.**
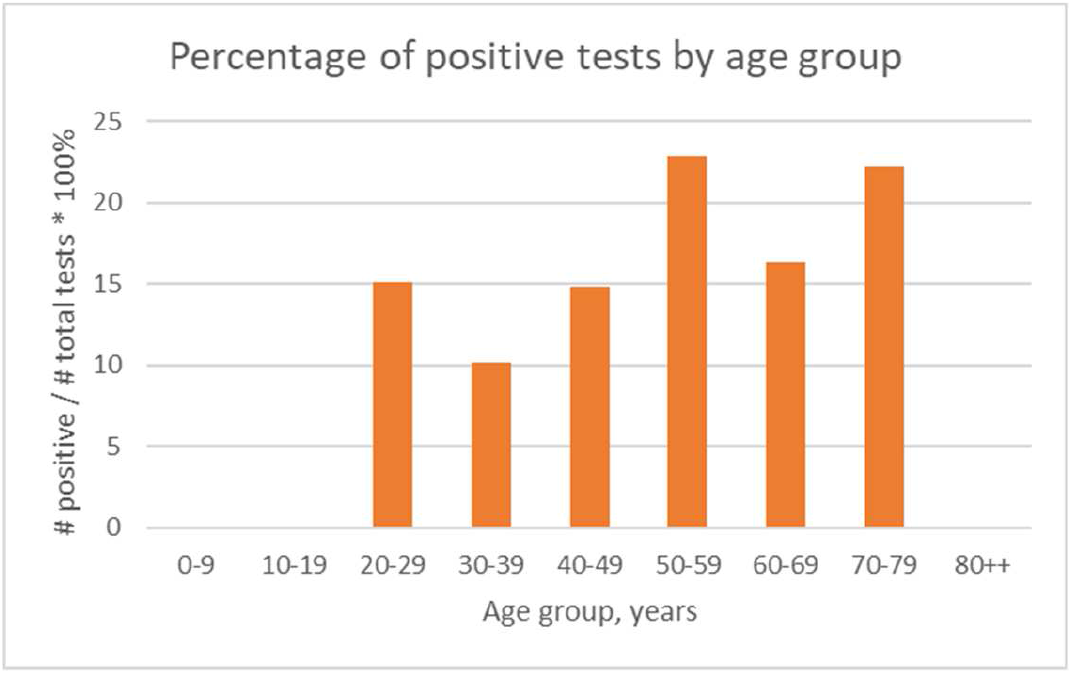
Percentage of positive tests by age group, obtained from Hospital X on May 05, 2020. Younger age groups have lower infection probability than older age groups.

## Conclusion

The ASQ model gives a much lower estimate on peak number of infected cases by 65.2% (3.46% vs 9.95%) compared with age-stratification alone; and by 75.2% (3.46% vs 13.99%) compared with Q-SEIR alone. We think this is because the median age of the Quezon City population is at a distance younger from the most vulnerable age group.

This method is part of a toolkit that forecasts demand on healthcare resources. More information on this toolkit can be found on https://sites.google.eom/a/dcs.upd.edu.ph/sms/simulating-the-dynamics-of-covid-19. Local government units and public officials may contact the corresponding author to request a demonstration. Improvements on this model may include nonlinear incidence and treatment rates [4] to incorporate social awareness, physical distancing and quality of patient treatment.

## Data Availability

Data available on request due to privacy/ethical restrictions.

http://ncovtracker.doh.gov.ph/

## Acknowledgements

We would like to thank the College of Engineering, University of the Philippines Diliman for supporting us in this research.

## Notes

### Competing Interest Statement

The authors have declared no competing interest.

### Funding Statement

The author(s) received no specific funding for this work.

